# Emulated Clinical Trials from Longitudinal Real-World Data Efficiently Identify Candidates for Neurological Disease Modification: Examples from Parkinson’s Disease

**DOI:** 10.1101/2020.08.11.20171447

**Authors:** Daphna Laifenfeld, Chen Yanover, Michal Ozery-Flato, Oded Shaham, Michal Rozen-Zvi, Nirit Lev, Yaara Goldschmidt, Iris Grossman

**Affiliations:** Formerly Teva Pharmaceuticals Research & Development, Petach Tikva, Israel; Formerly IBM Research – Haifa, Israel; IBM Research – Haifa, Israel; Faculty of Medicine, The Hebrew University, Jerusalem, Israel

**Keywords:** Real World Data, Clinical Evidence, Parkinson’s disease, Artificial Intelligence, Causal Inference, Rasagiline, Zolpidem, Disease Modifying Therapeutics, Repurposing

## Abstract

Real-world healthcare data hold the potential to identify therapeutic solutions for progressive diseases by efficiently pinpointing safe and efficacious repurposing drug candidates. This approach circumvents key early clinical development challenges, particularly relevant for neurological diseases, concordant with the vision of the 21^st^Century Cures Act. However, to-date, these data have been utilized mainly for confirmatory purposes rather than as drug discovery engines. Here, we demonstrate the usefulness of real-world data in identifying drug repurposing candidates for disease-modifying effects, specifically candidate marketed drugs that exhibit beneficial effects on Parkinson’s disease (PD) progression. We performed an observational study in cohorts of ascertained PD patients extracted from two large medical databases, Explorys SuperMart (N=88,867) and IBM MarketScan Research Databases (N=106,395); and applied two conceptually different, well-established causal inference methods to estimate the effect of hundreds of drugs on delaying dementia onset as a proxy for slowing PD progression. Using this approach, we identified two drugs that manifested significant beneficial effects on PD progression in both datasets: rasagiline, narrowly indicated for PD motor symptoms; and zolpidem, a psycholeptic. Each confers its effects through distinct mechanisms, which we explored via a comparison of estimated effects within the drug classification ontology. We conclude that analysis of observational healthcare data, emulating otherwise costly, large, and lengthy clinical trials, can highlight promising repurposing candidates, to be validated in prospective registration trials, for common, late-onset progressive diseases for which disease-modifying therapeutic solutions are scarce.

## Introduction

Repurposing of marketed drugs is an increasingly attractive prospect for drug developers and patients alike, given the ever-increasing costs of *de novo* drug development (Ashburn and Thor, 2004). The rationale underlying the practice of drug repurposing is supported by the demonstration, in a multitude of disease areas, of a drug’s mechanistic and clinical utility for multiple indications, ranging from migraine to autoimmune diseases (Xiao et al., 2008; Yong and D’Cruz, 2008; Cha et al., 2018). While the majority of repurposed drugs have been identified through serendipity, recent years have witnessed a growth in systematic efforts to identify new indications for existing drugs. These efforts include experimental screening approaches (Buckley et al., 2010; Deshmukh et al., 2013; Najm et al., 2015) and *in silico* approaches in which existing data are used to discover repurposing candidates [see (Cha et al., 2018) for in depth review of these methods]. Yet, key challenges in translating repurposing ideas into clinical application have hampered progress along this otherwise promising avenue.

Assessing the efficacy of a drug for any indication requires a series of independent analyses reporting data from humans treated with said drug, traditionally acquired through clinical trials. In the last decade, new opportunities have emerged for acquiring clinical evidence in manners complementing clinical trials, with the growing availability of real**-**world data (RWD), specifically electronic health records (EHRs) and medical insurance claims data, together with the advent of state-of-the-art computational methodologies. EHRs record multiple health-related data types over time, including drug prescriptions, lab test results of varying nature, physician visits, and symptomology, allowing the relationships between these different features to be assessed. Medical insurance claims data, another form of health related RWD, capture complementary and partially overlapping information, including medical billing claims, enabling research of hospitalizations, doctor’s visits, drug prescription and purchasing, and clinical utilization. In the context of drug repurposing, there have been isolated attempts to use RWD in a confirmatory capacity, to support clinical incidental findings. For example, EHRs have been used to demonstrate an association between metformin and decreased cancer mortality (Xu et al., 2014), and combined EHRs and claims data have been used to support the protective potential of L-DOPA against age-related macular degeneration (AMD) (Brilliant et al., 2016). Here, we propose a novel approach in which, for the first time, retrospective RWD is used to “industrialize serendipity”. We therefore systematically emulate Phase IIb studies for all concomitant medications used in a disease (for other than disease modifying purposes), in order to identify potential unexpected beneficial effects. Further, investigating the effects of related drugs, e.g., sharing target profile or mechanism of action, allows the extraction of mechanistic explanations for drug effect. These effects, once validated in multiple independent sources of RWD, provide robust evidence on drug efficacy, tolerability, and safety, as well as mechanistic insight on disease modification. It is therefore envisaged that drug candidates identified in this manner will leapfrog into the registration trial phase, confirming aims stated in the USA’s 21^st^ Century Cures Act (21st Century Cures Act, Pub. L. No. 114-255, 2016), and extending the European Medicines Agency (EMA) current use of RWD as an external control arm in rare disease clinical trials (Cave et al., 2019).

The complex nature and organ-inaccessibility of diseases related to the central nervous system (CNS) render them particularly attractive for an RWD-based approach of drug repurposing. For most CNS disorders, our understanding of pathophysiology and underlying etiology is still limited, resulting in poor availability of appropriate, mechanistically relevant, animal models. Furthermore, clinical trials testing disease-modifying agents require lengthy and large studies, burdening the patient population and incurring high costs of development. Together, these limitations constrain the ability of field experts to rationally design drugs that target these devastating diseases. Thus, using RWD to robustly explore the relationship between various drugs and co-morbidities for which they are not prescribed can help mitigate the risk of lack of predictive animal models, alongside the lengthy clinical studies required to determine outcome in the human setting. An example of such an approach is described in Mittal et al. (2017). The authors used the Norwegian Prescription Database to demonstrate that individuals prescribed salbutamol (Beta2-adrenoceptor agonist) had a lower incidence of Parkinson’s disease (PD), while those prescribed propranolol (Beta2-antagonist) exhibited higher PD incidence. However, investigation of disease progression or severity was not pursued.

PD is one of the most common neurodegenerative disorders, affecting 1 to 2 in 1,000 individuals worldwide and 1% of the population above 60 years of age (Tysnes and Storstein, 2017). To-date, no disease-modifying agents are approved for PD (Lang and Espay, 2018), highlighting the need and potential for novel approaches utilizing RWD to bring new therapies to late development stages, and thus quickly and effectively to PD patients. One of the hallmark clinical pathologies of PD progression is PD dementia (PDD) (Hely et al., 2008). An estimated 50-80% of PD patients experience dementia as their disease progresses, typically within 10 years of disease onset. It is therefore imperative to identify effective disease-modifying therapeutic agents (Aarsland and Kurz, 2010; Meireles and Massano, 2012). In this study, we used, for the first time, RWD from both EHRs and claims data to identify drugs associated with decrease in progression into PDD, as candidates for disease modification of PD. We applied a novel analytical framework of multiple, hierarchical “emulated PhIIb clinical trials”, an approach that inherently proposes mechanistic rationale for these drugs.

## Methods

### Study design

We used the drug repurposing framework (Ozery-Flato et al., 2020), emulating a PhIIb randomized controlled trial (RCT) for each candidate drug, combining subject matter expertise with data-driven analysis, and applying a stringent correction for multiple hypotheses. Specifically, each emulated RCT compared PD patients who initiated treatment with either the studied drug (treatment cohort) or an alternative drug (control cohort). We follow the target trial emulation protocol described by Hernán and Robins (2016), which includes the following steps: define the study eligibility criteria; assign patients to treatment and control cohorts; list and extract a comprehensive set of per-patient baseline covariates; list and extract follow-up disease-related outcome measures; and, finally, use causal inference methodologies (Hernan and Robins, 2020) to retrospectively estimate drug effects on disease outcomes, correcting for confounding and selection biases. We next elaborate on each of these protocol components.

### Data Sources

We analyzed two individual-level, de-identified medical databases. The IBM Explorys Therapeutic Dataset (“Explorys”; freeze date: August 2017) includes medical data of >60 million patients, pooled from multiple healthcare systems, primarily clinical EHRs. The IBM MarketScan Research Databases (“MarketScan”; freeze date: mid 2016) contain healthcare claims information from employers, health plans, hospitals, Medicare Supplemental insurance plans, and Medicaid programs, for ∼120 million enrollees between 2011 and 2015.

### Eligibility Criteria

Patients were included in the PD cohort based primarily on diagnosis codes (Table S1), using the International Classification of Diseases (ICD) system (ICD-9 and ICD-10). We required a repeated PD diagnosis on two distinct dates and excluded patients with secondary parkinsonism or non-PD degenerative disorders. We further excluded early-onset (age <55 years) PD, and patients with metastatic tumors or those ineligible for prescription drugs through their medical insurance plans. *PD initial date* was set to the earliest date of first PD diagnosis or a levodopa (an approved symptomatic therapy for PD, compensating for the depleted supply of endogenous dopamine) prescription within the year preceding the first diagnosis of the disease. Since PD is likely present latently before the first diagnostic or prescription record, we retracted the disease date by additional six months. We only included patients whose PD initial date preceded the date of treatment assignment, which we termed *index date*. To ensure accurate characterization of a patient’s clinical state, we required data history of at least one year prior to the index date. Finally, we excluded from the control cohort patients who were prescribed the trial drug.

### Treatment Assignment

For both treatment and control cohorts, we demanded the assigned treatment to have at least two prescriptions 30+ days apart. To avoid confounding by indication, we considered alternative drugs that shared the same (or similar) therapeutic class. Specifically, we first compared each studied drug to drugs taken from its second level Anatomical Therapeutic Chemical (ATC) (World Health Organization, 2020) class. Then, for each drug candidate showing a significant beneficial effect across the two databases, we expanded the analysis to control cohorts corresponding to ATC classes of all levels.

### Outcomes and Confounders

The primary endpoint was newly diagnosed dementia during a follow-up period of two years (starting at the index date) as proxy for PD progression;patients with a dementia-related diagnosis at baseline were excluded. Other supporting endpoints considered were falls and psychosis prevalence (see Table S3 for defining ICD codes). We extracted hundreds of pre-treatment patient characteristics (Ozery-Flato et al., 2017) (throughout the one year preceding the index date), covering those identified by a subject matter expert as potentially associated with confounding or selection bias. These included demographic attributes, co-morbidities, PD related diagnoses, PD related drugs, non-PD drugs, healthcare services utilization and socioeconomics parameters (Table 1).The extracted covariates provide a multifaceted view of a patient’s PD status at the index date, as manifested in the medical records of the patient prior to RCT initiation.

**Table 1.** Pre-treatment patient characteristics (considered as potential confounders).

| Characteristic | Description |
| --- | --- |
| **Characteristics related to PD severity** |  |
| PD related diagnoses (Table S3) | Dementia, measuring progression along the cognitive axis; falls, as a proxy to advanced motor impairment and dyskinesia; and psychosis, measuring progression along the behavioral axis. |
| PD related drugs | Indicators for prescriptions of PD indicated drugs (Table S2) during the baseline period, excluding the index-date; indicators for anti-cholinergic (ATC N04A), dopaminergic (ATC N04B), anti-psychotics (ATC N05A) and anti-dementia (ATC N06D) drugs. |
| **General characteristics** |  |
| Age | At index date |
| Gender | Male, Female, unknown |
| Socioeconomics | Indicators for commercial, Medicare-Supplemental, and Medicaid insurance (MarketScan only). |
| Co-Morbidities | Charlson's comorbidity index (Charlson et al., 1987); an indicator per each of its underlying comorbidity category; and an indicator per each diagnosis class, using the Clinical Classification Software (Clinical Classifications Software (CCS) 2015) |
| Other drugs | Indicators for Level 2 ATC class of all prescribed drugs. |
| Healthcare services utilization | Counts of distinct visit dates per provider place and type (MarketScan only); total days of admission per encounter type (Explorys only); indicator for index date drug prescription during inpatient hospitalization (Explorys only). |

### Statistical Analysis

The effect of the trial drug on disease progression was evaluated as the difference between the expected prevalence of the outcome event for drug-treated patients and that in control patients during a complete follow-up period. Briefly, we corrected for potential confounding and selection biases, using two conceptually different causal inference approaches: (i) *balancing weights*, via Inverse Probability Weighting (IPW) (Austin, 2011), which reweighs patients to emulate random treatment assignment and uninformative censoring; and (ii) *outcome model*, using standardization (Hernan and Robins, 2020) to predict counterfactual outcomes. We considered a confounder as balanced if the standardized mean difference between (weighted) treatment and control cohorts was below 0.2. We analyzed Explorys and MarketScan separately and focus here on the overlapping, statistically significant, candidates. This stringent approach bypasses the need to arbitrarily set aside one database as “confirmatory” and it extends more straightforwardly to >2 data resources. Finally, we used Benjamini and Hochberg’s (1995) method to correct for multiple hypothesis testing and considered adjusted P-values ≤ 0.05 as statistically significant. For a full description of the RWD-based drug repurposing framework see our methodological paper (Ozery-Flato et al., 2020). Ground truth effects (that is, RCT-validated) are typically unavailable for drug repurposing candidates; notably, however, the estimated effects showed significant correlation across different algorithms and data sources (adjusted P-value < 0.05 for all comparisons across outcomes, databases, and causal inference algorithms), attesting to the robustness of the framework.

## Results

We first extracted cohorts of late-onset PD patients comprising approximately 106,000 and 89,000 patients in MarketScan and Explorys, respectively, representing 0.09% and 0.15% of the total databases and consistent with recent epidemiological surveys (Tysnes and Storstein, 2017). Key characteristics of these separate cohorts (Table 2) exhibit high similarity in the average and range of age at PD initial date, the percentage of women, the fraction of patients with public insurance, and the baseline Charlson comorbidity index (Charlson et al., 1987). Notable dissimilarities between the two cohorts include the average total patient time in database, which was more than double in Explorys compared to MarketScan (Table 2). This dissimilarity stems from the different timespan covered in general by the two databases (average total patient timeline of 4.7±17.4 years in Explorys vs. 2.2±1.6 years in MarketScan). We note that for most patients, PD initial date does not correspond to disease onset, which is often unknown and may precede the observed time period. Nevertheless, ascertainment of PD status at the index date can be reliably characterized by triangulation of the patient’s history of PD-related diagnoses, PD medications, and healthcare utilization. In our two PD cohorts, dementia was the most prevalent PD-related diagnosis (7.6-9.2%) at PD initial date, followed by fall and psychosis (1.4-4.1%).

**Table 2.** PD cohort characteristics.

|  | MarketScan | Explorys |
| --- | --- | --- |
| **Patient count** | 106,395 | 88,867 |
| **Patient timeline [years]** |  |  |
| **Total<sup>a</sup>** | 3.0 (1.6) [1.7; 3.0; 5.0] | 7.6 (5.7) [2.9; 6.5; 11.6] |
| **Before PD initial date<sup>a</sup>** | 0.7 (1.1) [0.0; 0.0; 1.0] | 4.3 (5.0) [0.0; 2.4; 7.3] |
| **After PD initial date<sup>a</sup>** | 2.3 (1.4) [1.1; 2.0; 3.2] | 3.3 (2.7) [1.2; 2.7; 4.7] |
| **No. of unique prescribed drugs<sup>a</sup>** | 14.5 (10.4) [7.0; 13.0; 20.0] | 13.1 (18.7)<br>[0.0; 5.0; 19.0] |
| **Insurance** |  |  |
| **Medicare, Medicaid, other public** | 90280 (84.9%) | 65146 (73.3%) |
| **Commercial, private only** | 16115 (15.1%) | 10810 (12.2%) |
| **Other or unknown** | 0% | 12911 (14.5%) |
| **Baseline characteristics (during ≤1 year before PD initial date)** |  |  |
| **Age at PD initial date<sup>a</sup>** | 74.8 (10.0) [66.2; 75.6; 82.7] | 74.3 (8.1) [68.6; 75.3; 80.7] |
| **Women** | 49,693 (46.7%) | 37,958 (42.7%) |
| **Charlson's Comorbidity Index<sup>a</sup>** | 0.7 (1.6) [0.0; 0.0; 1.0] | 0.6 (1.3) [0.0; 0.0; 1.0] |
| **PD-related diagnoses** |  |  |
| **Falls** | 1746 (1.6%) | 3604 (4.1%) |
| **Psychosis** | 2368 (2.2%) | 1209 (1.4%) |
| **Dementia** | 9761 (9.2%) | 6716 (7.6%) |
| **Follow-up characteristics (during ≤2 years following PD initial date)** |  |  |
| **Dementia<sup>b</sup>** | 43806, 45% (41.2%) | 25446, 32% (28.6%) |
| **Charlson's Comorbidity Index<sup>a</sup>** | 2.8 (2.8) [1.0; 2.0; 4.0] | 1.8 (2.4) [0.0; 1.0; 3.0] |

<sup>a</sup> Mean, standard deviation (in parentheses), and the first, second (median), and third quartile (in brackets).
<sup>b</sup> Population-level follow-up prevalence of dementia corresponds to the Kaplan-Meier estimator, which adjust for censoring, with the non-adjusted prevalence given in parentheses.

Overall, we tested all (n=218) drugs whose treatment and control cohorts had each at least 100 PD patients in both the MarketScan and Explorys. We used this lower bound since many Phase IIb and III clinical trials, including those pursued in neurological indications, find 100 or less patients per arm to be satisfactory. Of these, we were able to balance all observed confounding biases between the treatment and control cohorts (using IPW, see Methods) for 205 drugs (94%). Consequently, for each such drug we emulated a two-year RCT, estimating its effect on the population-level prevalence of newly diagnosed dementia, in comparison to the level-2 Anatomical Therapeutic Chemical (ATC) control cohort. Using two independent causal inference methods, outcome model and balancing weights, our analysis identified, in both data sources, two candidate drugs estimated to significantly reduce dementia prevalence: rasagiline and zolpidem (see cohort characteristics in Tables S4-S7).

Details of the emulated RCTs, estimating the effect of these drugs compared to their corresponding control ATC level-2 class, are shown in Tables 3,4, S8 and S9. Figure 1 shows the prevalence of newly diagnosed dementia in the treatment and control cohorts throughout the follow-up period. Consistently, rasagiline is estimated to decrease the prevalence of newly diagnosed dementia during a follow-up period of two years by 7-9%, compared to symptomatic PD drugs. Similarly, Zolpidem, compared to the class psycholeptics drugs, reduces dementia prevalence by 8-12%. Moreover, for both rasagiline and zolpidem, drug effect increases as a function of treatment duration (Figure 1). We emphasize that in these emulated RCTs, as well as the ones discussed below, the causal methodology we applied successfully balanced the treatment and control cohorts with respect to all hypothesized confounders (Table 1), suggesting that important characteristics of these cohorts, including age and proxies of disease stage, are now similar.

**Table 3.**
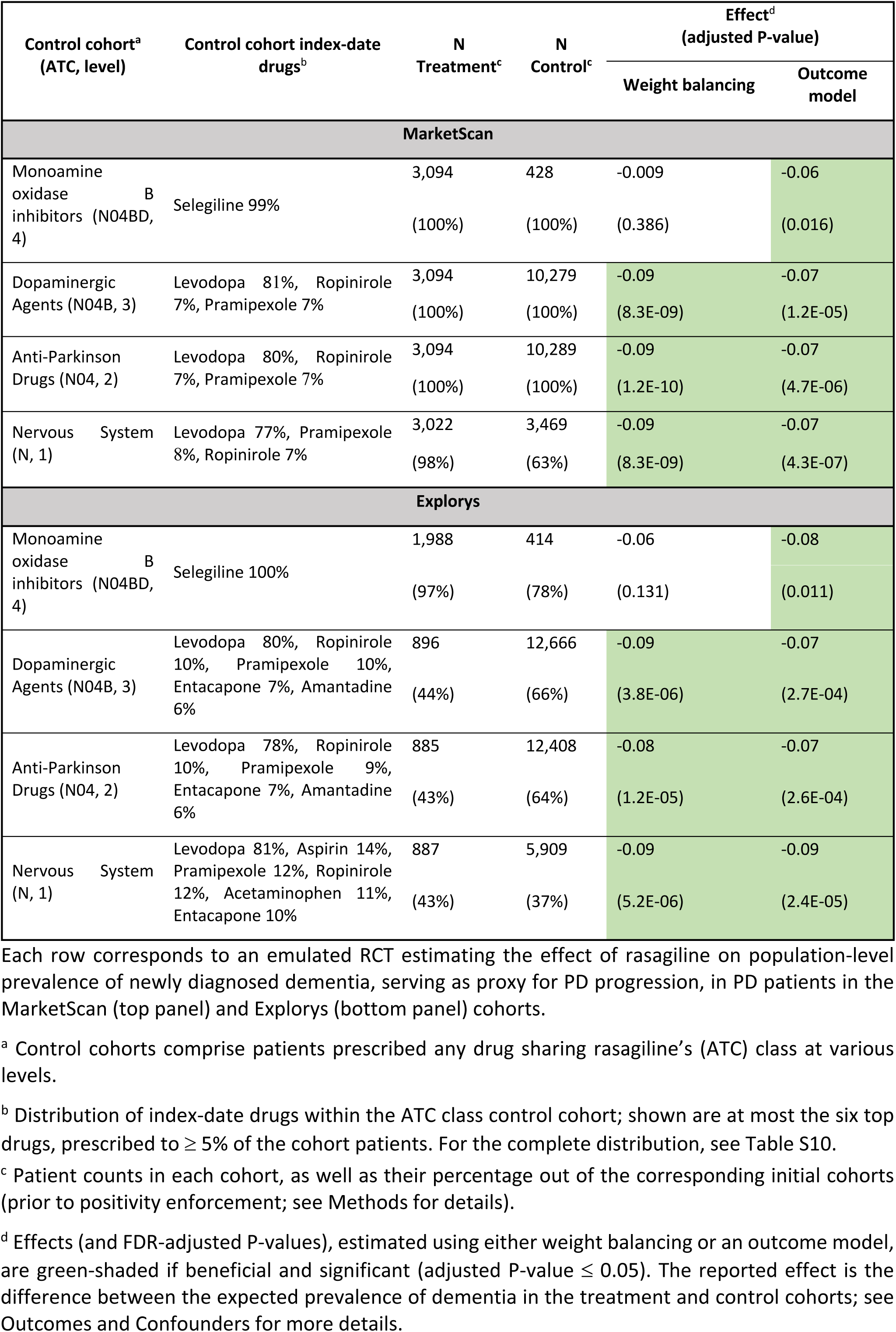
Rasagiline significantly attenuates PD progression.

**Table 4.**
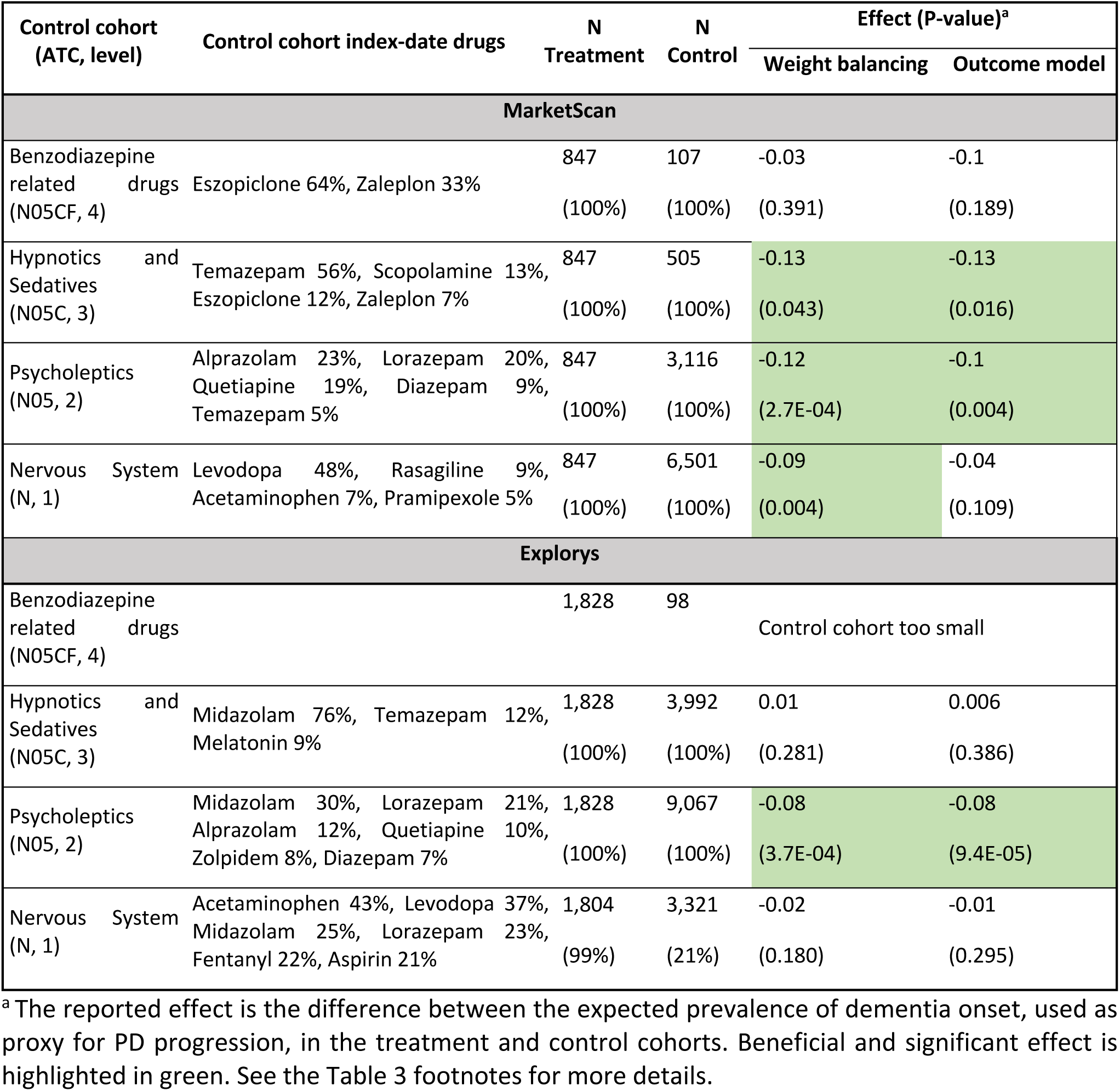
Zolpidem significantly attenuates PD progression.

**Figure 1:**
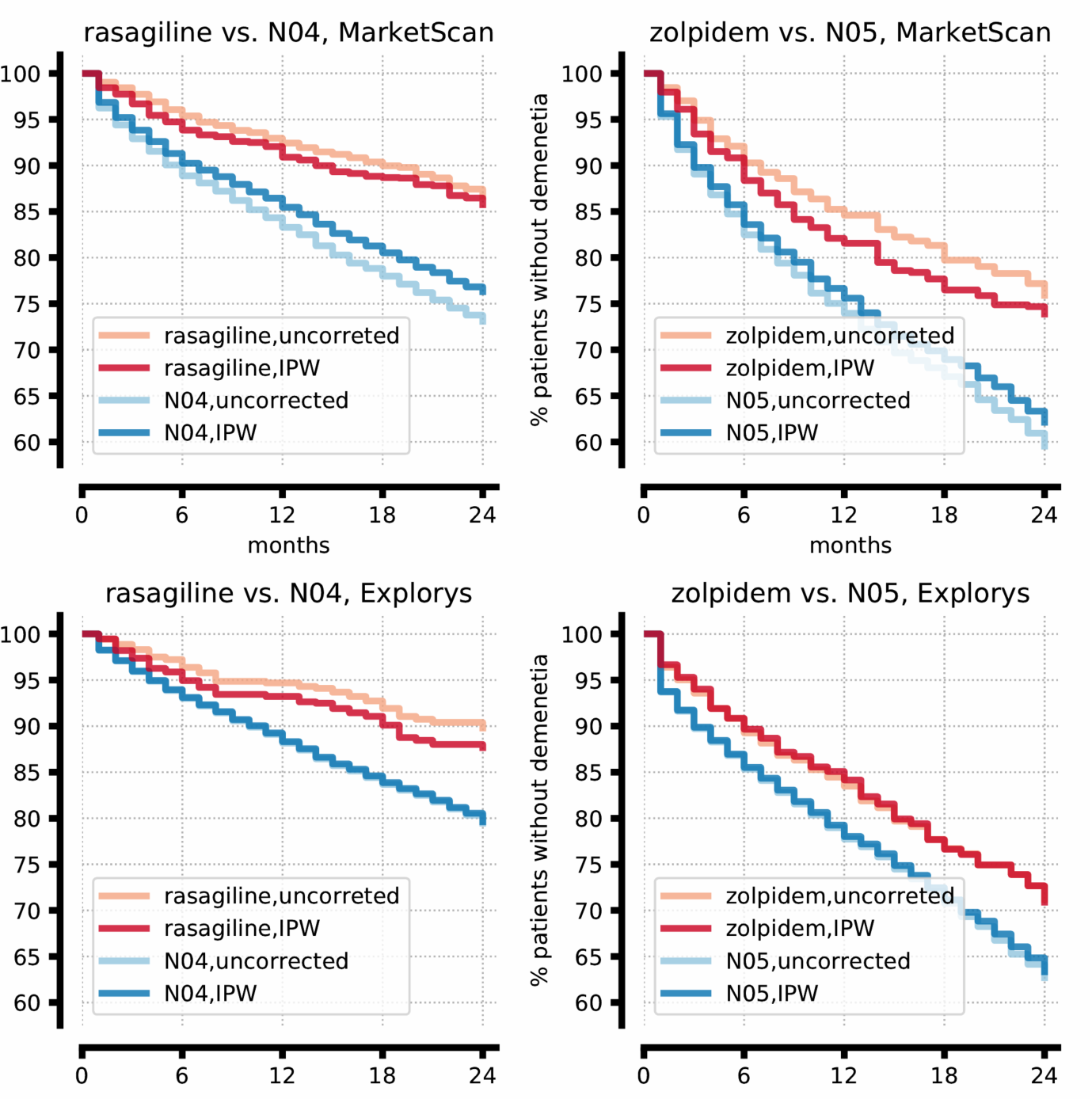
Rasagiline and Zolpidem significantly delay the onset of dementia in PD patients in two independent datasets. Kaplan-Meier plots comparing the prevalence of newly diagnosed dementia in the treatment and control cohorts, corrected with inverse probability weighting (IPW, dark color), or uncorrected (light color). Red and blue lines show the expected percentage of patients not yet diagnosed with dementia at each time point among the patients who take the drug and among the patients who take other ATC level 2 drugs (N04: symptomatic PD drugs; N05: Psycholeptics), respectively. The difference between each pair of red and blue lines correspond to the expected effect of the drug.

Next, we expanded the analysis to consider all four ATC levels that include each drug, corresponding to anatomical main group (level 1), therapeutic subgroup (level 2), pharmacological subgroup (level 3), and chemical subgroup (level 4). Specifically, we compared each drug against all its encompassing ATC classes and additionally, each encompassing ATC class against all upper-level classes in the ATC hierarchy. The resulting set of RCTs estimates the effect of a target drug against drugs sharing its mechanism of action (MoA; e.g., rasagiline versus other monoamine oxidase B inhibitors, ATC class N04BD), as well as drugs conferring different MoAs (e.g., monoamine oxidase B inhibitors, N04BD, versus other dopaminergic agents, N04B), thus testing a set of related mechanistic hypotheses. This can also be viewed as sensitivity analyses for the effect of a target drug. Table S11 shows the complete results of these emulated RCTs.

ATC level-4 class N04BD, monoamine oxidase (MAO) B inhibitors, included only two drugs: rasagiline and selegiline. Therefore, the rasagiline versus N04BD emulated trial is essentially a head-to-head comparison between these two drugs. The results of the emulated trials in both MarketScan and Explorys suggest that the use of rasagiline reduces the prevalence of dementia compared to selegiline (Table 3; estimations using outcome model are significant). When compared to higher level ATC classes – specifically, dopaminergic agents, symptomatic PD drugs, and nervous system medications – all dominated by levodopa (77-82% of first prescriptions), rasagiline is estimated to significantly decrease dementia prevalence by 5-9% in both databases, using either causal inference approach (Table 3, and Table S8). We also estimated the effect of rasagiline on the prevalence of falls and psychosis: In MarketScan, rasagiline is estimated, by both causal inference algorithms, to decrease the population prevalence of falls compared to all its encompassing ATC classes; in Explorys, rasagiline is estimated to have a beneficial effect on the prevalence of psychosis (but only a subset of these estimands were significant).

Zolpidem was estimated to have significant and beneficial effects on the prevalence of dementia only in comparison to its level-2 ATC class, psycholeptics (Table 4, and Table S9). The analysis in MarketScan suggests that zolpidem has a beneficial effect compared to other hypnotics and sedatives (N05C), but the different composition of the N05C control cohort in Explorys (dominated by midazolam) hinders conclusive results. Zolpidem was also estimated to have beneficial effects on the prevalence of falls and psychosis, compared to psycholeptics, but these effects were not significant.

## Discussion

The present study used both EHRs and insurance claims data to assess the effects of hundreds of concomitant drugs on the emergence of PD-associated dementia as one of the more common hallmarks of PD progression. Only those drugs for which a statistically significant effect was found independently in both EHR and claims data were further considered for their repurposing potential. Given the different nature of the data collected with each health data source and stringent statistical approach, the resultant repurposing candidates have a high likelihood of success in a Phase III prospective study. Our analysis unraveled therapeutic benefits of two drugs in decreasing the population-level incidence of PDD, representing slowing of PD disease progression. Thus, long-term treatment (24 months) with rasagiline, a MAO-B inhibitor narrowly indicated for PD motor symptoms, or with zolpidem, a gamma-aminobutyric acid (GABA)-A receptor modulator indicated for insomnia, is strongly associated with decreased PDD incidence in two separate large cohorts (N=195,262 in total). Indeed, the mechanistic, and at times clinical, support for the identified associations, as described below, not only provides support for the approach in identifying new drug repurposing candidates, but also a vehicle to bolster otherwise ambiguous results from RCTs. We note that in a similar analysis we also found azithromycin and valsartan to significantly decrease the prevalence of falls and psychosis, respectively, in PD patients but without significantly reducing the rate of dementia onset; discussion of these drug repurposing candidates is beyond the scope of the current publication.

Cognitive impairment is highly prevalent in patients with progressive stages of PD and is associated with adverse health outcomes and increased mortality (Bäckström et al., 2018). Cognitive deficits vary in quality and severity in different stages of disease progression in PD, ranging from subjective cognitive decline to mild cognitive impairment and to subsequent PDD. The latter is defined as acquired objective cognitive impairment in multiple domains, including attention, memory, executive and visuospatial ability (Emre et al., 2007), and results in adverse alteration of activities of daily life (American Psychiatric Association, 2013). In a study of 224 Norwegian PD patients (Aarsland et al., 2003), for whom disease duration was 9 years on average, the estimated 4-year and 8-year prevalence of dementia was 51.6 % and 78.2% respectively. In another study that followed 136 newly diagnosed PD patients for 20 years (Hely et al., 2008), dementia was present in 83% of 20-year survivors. A single choline esterase inhibitor, rivastigmine, is approved by the US Food and Drug Administration (FDA) for the treatment of PDD, with modest efficacy (Meng et al., 2018) resulting in a significant unmet medical need for additional pro-cognitive therapies (Green et al., 2019).

Our finding that rasagiline slows PD progression is consistent with mechanistic evidence and extends prior clinical data. Clinical trials of rasagiline in PD patients implied possible disease-modifying effects, albeit inconclusively. Indeed, none of the studies reported to-date had the statistical power to support or refute slowing the progression of the disease. The largest study to assess disease-modifying effects of rasagiline was ADAGIO (Olanow et al., 2009), which failed to demonstrate a dose-dependent effect on the Unified Parkinson’s Disease Rating Scale (UPDRS) scores. This failure may be partly due to insufficient statistical power: the total number of participants in the ADAGIO study was N=1,176, much smaller than in our study (N=13,562 in Explorys; N=13,373 in MarketScan; See Table 3). Additionally, the ADAGIO study did not directly assess effects of rasagiline on cognition. Several recent studies addressed this hypothesis more directly, but were small (N=34-151) and short (3-6 months), yielding mixed results (Hanagasi et al., 2011; Frakey and Friedman, 2017). A larger study (N=289 completers) assessed similar, but distinct effects of rasagiline as add-on therapy (Hauser et al., 2014), reporting a statistically significant improvement when added to dopamine agonist therapy over 18 months of therapy. Importantly, many of the prior reports sought to demonstrate disease prevention/protection in as-yet-to-be-diagnosed patients, while we studied patients with confirmed PD diagnosis. Due to this important distinction, it can be expected that the class and specific agents reported, e.g., by Mittal et al. (2017), to decrease (or increase) PD incidence did not show, in our analysis, similar effects. Overall, inadequate power and diverse study designs hampered conclusive therapeutic interpretation of the role of rasagiline, and the monoamine B class, as PD disease modifiers. Indeed, our approach directly resolved these shortcomings, dramatically increasing sample size and follow-up duration by virtue of the use of RWD, facilitating the discovery of rasagiline’s robust and consistent disease-modifying effects. Importantly, our analysis of proxy parameters supports the beneficial effects of rasagiline on PD progression beyond PDD, as reflected by a decrease in the population prevalence of falls and the trend reduction of psychosis (data not shown).

Mechanistically, rasagiline has been suggested to have neuroprotective effects mediated by its ability to prevent mitochondrial permeability transition (Naoi and Maruyama, 2009). In addition, rasagiline induces anti-apoptotic pro-survival proteins, Bcl-2 and glial cell-line derived neurotrophic factor (GDNF) and increases expression of genes coding for mitochondrial energy synthesis, inhibitors of apoptosis, and the ubiquitin-proteasome system. Finally, systemic administration of selegiline and rasagiline increases neurotrophic factors in cerebrospinal fluid of PD patients and non-human primates (Naoi et al., 2007).

The association between zolpidem, a non-benzodiazepine hypnotic drug used for the treatment of sleeping disorders, and decreased PDD incidence identified herein is a novel finding. In fact, a single prior report published more than two decades ago speculated that zolpidem would not be efficacious for PD, based on the limited clinical experience with the drug at the time, without specific consideration for cognition (Lavoisy and Marsac, 1997). However, recent publications demonstrate Zolpidem’s ability to treat a large variety of neurologic disorders, most often related to movement disorders and disorders of consciousness, and suggest zolpidem induces transient effects on UPDRS (Bomalaski et al., 2017). Of note, several cross-sectional reports have raised concerns for increased risk of reversible dementia or Alzheimer’s diseases in the general population when exposed to zolpidem (Shih et al., 2015; Lee et al., 2018). However, these reports considered only a handful of potential confounding biases, and applied regression-based methods, which unlike IPW, do not allow one to determine whether treatment and control biases were successfully eliminated (Austin, 2011). Furthermore, neither report assessed impact on specific patient subsets, such as those diagnosed with PD. Indeed, a proof-of-concept clinical study is currently recruiting subjects in order to assess the benefits of low-dose zolpidem in late-stage PD (NCT03621046), supporting the findings reported herein. Yet again, the limited sample size (N=28) in the recruiting study, together with the inclusion of cognition as a secondary (rather than primary) endpoint both pose a high risk for insufficient power and thus inconclusive results. Finally, latest literature reports on beneficial effects of zolpidem on renal damage and akinesia (Bortoli et al., 2019) support a high benefit-risk profile of repurposing zolpidem for slowing or reversal of PD.

Mechanistically, a structural relationship between the antioxidant melatonin and zolpidem suggests possible direct antioxidant and neuroprotective properties of zolpidem. Garcia-Santos et al (2004) demonstrated that zolpidem prevented induced lipid peroxidation in rat liver and brain homogenates, showing antioxidant properties similar to melatonin. Bortoli et al. (2019) investigated *in silico* the antioxidant potential of zolpidem and identified it as an efficient radical scavenger similar to melatonin and Trolox. Although the mechanisms involved in the pathogenesis and progression of PD are not fully understood, there is overwhelming evidence that oxidative stress plays an important role in dopaminergic neuronal degeneration. Since the maintenance of reduction-oxidation reaction potential is an important determinant of neuronal survival (Puspita et al., 2017), its disruption ultimately leads to cell death. Accumulating evidence from patients and disease models indicate that oxidative and nitrative damage to key cellular components is important in the pathogenesis of PD progression (Vera et al., 2013). Oxidative stress plays an important role in dopaminergic neuronal degeneration, triggering a cascade of events, including mitochondrial dysfunction, impairment of nuclear and mitochondrial DNA, and neuroinflammation, which in turn cause more reactive-oxygen species (ROS) production (Guo et al., 2018). Genetic forms of PD, including those caused by mutations in *PARK7, PINK1, PRKN, SNCA* and *LRRK2*, also demonstrate the fundamental role that mitochondrial function plays in disease etiology (Vera et al., 2013). Thus, the protective effects of zolpidem on the development of dementia could be explained by the antioxidant and neuroprotective capacities of the drug.

In a preliminary method development study (Ozery-Flato et al., 2020), we validated the drug repurposing framework used here. We demonstrated that treatment effects estimated across different data sources and causal methodologies showed a high degree of agreement (P-value < 0.05 for all comparisons). Yet, the retrospective design of the study, combined with the use of RWD, introduces some limitations. Specifically, identifying phenotype cohorts based on ICD codes is likely to be incomplete (sensitivity <1) and noisy (positive predictive value, PPV < 1). None withstanding, it is considered a fairly accurate and practical approach to “rule in” patients with PD (Noyes et al., 2007). Corroborating these assignments by medical history and drug prescriptions further substantiates patients’ eligibility. Additionally, proxies with reliable representation in the data are required to emulate the endpoints in prospective clinical trials and need to be further assessed and refined in a controlled clinical environment (Shivade et al., 2014). Still, automatic mapping of EHR data to phenotypes and medical concepts needed for clinical research has gained much attention, yielding multiple studies that demonstrate the increasing ability of machine learning and artificial intelligence to provide accurate solutions for this challenge (Hripcsak and Albers, 2013; Ho et al., 2014; Beaulieu-Jones and Greene, 2016; Lipton et al., 2017). Conversely, the mechanistic nature of the drug effects, and therefore potential utility in combination therapy for synergistic effects, require further assessment in a dedicated prospective study, consistent with the drug development paradigm. In addition, while RWD used in a retrospective manner enables the assessment of chronic processes, without the need for lengthy studies, they are bound by the length of follow-up data per individual. Finally, local healthcare practice may at times confound the analysis and requires in-depth understanding of such practices in data interpretation (Hersh et al., 2013).

Notwithstanding these limitations, discoveries stemming from RWD of large, well-characterized patient populations can provide valuable clues to effective mechanisms and existing medications that may be beneficial in slowing disease progression, or potentially preventing it altogether. In the realm of CNS-related diseases, the extensive follow-up integral to medical-record tracking presents a well-suited setting for investigating the effects of concomitant interventions. Our two-year follow-up period is longer compared to most PD clinical trials, including the ones discussed above, and can be further prolonged in Explorys (see timeline statistics in Table 2). The EMA has already employed RWD in lieu of control arms to support regulatory decisions either at authorization or for indication extension, in the context of rare, orphan diseases (Cave et al., 2019). Similarly, the 21st Century Cures Act (21st Century Cures Act, Pub. L. No. 114-255, 2016) requires that the FDA establishes a framework to evaluate the potential use of RWD in support of approval of new indications for approved drugs. In fact, successful examples are already being implemented (Baumfeld Andre et al., 2019). Accordingly, the FDA allotted $100 million to build an EHR database of 10 million people as a foundation for more robust postmarketing studies. The current study provides evidence in support of such uses for RWD, accelerating the availability of solutions for patients in need.

In conclusion, we demonstrated that emulating clinical trials based on observational healthcare data identifies promising repurposing drug candidates, efficiently relieving the societal burden of costly, large, and lengthy clinical trials. This approach is particularly relevant as a therapeutic discovery engine for common, late-onset progressive CNS diseases for which disease-modifying therapeutic solutions are scarce. As the PD population is heterogenous, refining the inclusion/exclusion criteria of the targeted sub-populations to focus on responder populations, compared to matched controls, will further increase the power of future analyses (Ozery-Flato et al., 2018). The two drugs identified herein, rasagiline and zolpidem, both hold great promise as disease-modifying agents for PD, in general, and specifically in addressing aspects of cognitive impairment in PD. Further, these cognitive benefits may extend to other neurodegenerative diseases. The ability to systematically compare effects between various drug classes, as well as within classes, in patients in real-world settings is a significant step in accelerating patients’ access to safe and efficacious therapies.

## Data Availability

The data analyzed in this paper is available in (i) IBM Explorys Therapeutic Dataset and (ii) IBM MarketScan Research Databases.
https://www.ibm.com/products/marketscan-research-databases
https://www.ibm.com/watson-health/about/explorys

https://www.ibm.com/products/marketscan-research-databases

## Supplementary Information

**Table S1**. ICD codes for PD cohort definition.

**Table S2**. PD-indicated drugs and their corresponding ATC class names.

**Table S3**. PD outcome definitions.

**Table S4**. Rasagiline cohort characteristics.

**Table S5**. Zolpidem cohort characteristics.

**Table S6**. N04 cohort characteristics.

**Table S7**. N05 cohort characteristics.

**Table S8**. Estimated effects on dementia onset for emulated RCTs involving rasagiline and its encompassing ATC classes.

**Table S9**. Estimated effects on dementia onset for emulated RCTs involving zolpidem and its encompassing ATC classes.

**Table S10**. Trial drugs distribution. (Trial_drugs.xlsx)

**Table S11**. Emulated RCTs outcomes and effects. (RCTs outcomes and effects.xlsx)

