## Supplementary Material for "Emulated Clinical Trials from Longitudinal Real-World Data Efficiently Identify Candidates for Neurological Disease Modification: Examples from Parkinson’s Disease"

**Table S1. ICD codes for PD cohort definition**

| Type | System | Code | Name |
|---|---|---|---|
| Inclusion | icd9 | 3320 | Paralysis agitans |
| | icd10 | G20 | Parkinson's disease |
| Exclusion | icd9 | 3316 | Corticobasal degeneration |
| | icd9 | 3321 | Secondary parkinsonism |
| | icd9 | 3330 | Other degenerative diseases of the basal ganglia |
| | icd10 | G21 | Secondary parkinsonism |
| | icd10 | G231 | Progressive supranuclear ophthalmoplegia [Steele-Richardson-Olszewski] |
| | icd10 | G3185 | Corticobasal degeneration |

**Table S2. PD-indicated drugs and their corresponding ATC class names.** The list below has been compiled by a domain expert based on the following sources: National Drug File – Reference Terminology (NDF-RT), Anatomical Therapeutic Chemical Classification System (ATC), and DrugBank ***(45)***.

| Drug | ATC class code(s) | ATC class name(s) |
|---|---|---|
| Amantadine | N04BB | Adamantane derivatives |
| Apomorphine | G04BE; N04BC | Drugs used in erectile dysfunction; dopamine agonists |
| Atropine | A03BA; S01FA | Belladonna alkaloids, tertiary amines; anticholinergics |
| Benztropine | N04AC | Ethers of tropine or tropine derivatives |
| Biperiden | N04AA | Tertiary amines |
| Bornaprine | N04AA | Tertiary amines |
| Bromocriptine | G02CB; N04BC | Prolactine inhibitors; dopamine agonists |
| Budipine | N04BX | Other dopaminergic agents |
| Cabergoline | G02CB; N04BC | Prolactine inhibitors; dopamine agonists |
| Carbidopa | N/A | N/A |
| Dexetimide | N04AA | Tertiary amines |
| Dihydroergocryptine | N04BC | Dopamine agonists |
| Entacapone | N04BX | Other dopaminergic agents |
| Hyoscyamine | A03BA | Belladonna alkaloids, tertiary amines |
| Levodopa | N04BA | Dopa and dopa derivatives |
| Methixene | N04AA | Tertiary amines |
| Orphenadrine | N04AB; M03BC | Ethers chemically close to antihistamines; ethers, chemically close to antihistamines |
| Pergolide | N04BC | Dopamine agonists |
| Piribedil | N04BC | Dopamine agonists |
| Pramipexole | N04BC | Dopamine agonists |
| Procyclidine | N04AA | Tertiary amines |
| Profenamine | N04AA | Tertiary amines |
| Rasagiline | N04BD | Monoamine oxidase B inhibitors |
| Ropinirole | N04BC | Dopamine agonists |
| Rotigotine | N04BC | Dopamine agonists |
| Selegiline | N04BD | Monoamine oxidase B inhibitors |
| Tolcapone | N04BX | Other dopaminergic agents |
| Trihexyphenidyl | N04AA | Tertiary amines |
| Tropatepine | N04AA | Tertiary amines |

**Table S3. PD outcome definitions.**

| Outcome | System | Code | Name |
|---|---|---|---|
| Fall | ICD9 | E88 | Accidental falls |
| | ICD10 | W0, W1 | Falls |
| Psychosis | ICD9 | 292 | Drug-induced mental disorders |
| | | 297 | Delusional disorders |
| | | 298 | Other nonorganic psychoses |
| | ICD10 | F2 | Schizophrenia, schizotypal and delusional disorders |
| Dementia | ICD9 | 294 | Persistent mental disorders due to conditions classified elsewhere |
| | | 331 | Other cerebral degenerations |
| | ICD10 | F01 | Vascular dementia |
| | | F02 | Dementia in other diseases classified elsewhere |
| | | F03 | Unspecified dementia |
| | | G30 | Alzheimer disease |

**Table S4. Rasagiline cohort characteristics.** Shown are the mean, standard deviation (in parentheses), and the first, second (median), and third quartile (in brackets).

| | MarketScan | Explorys |
|---|---|---|
| **Patient count** | 3,094 | 1,988 |
| **Patient timeline [years]** | | |
| **Total** | 4.1 (1.0) [3.0; 4.6; 5.0] | 11.2 (5.1) [7.0; 10.6; 14.8] |
| **Before index date** | 2.5 (1.0) [1.6; 2.4; 3.2] | 7.3 (4.8) [3.3; 6.3; 10.6] |
| **After index date** | 1.6 (1.0) [0.8; 1.5; 2.4] | 3.9 (2.5) [2.0; 3.5; 5.4] |
| **No. of unique prescribed drugs** | 15.6 (9.3) [9.0; 14.0; 21.0] | 20.7 (18.2) [7.0; 15.0; 29.0] |
| **Insurance** | | |
| **Medicare, Medicaid, other public** | 60% | 80% |
| **Commercial, private only** | 40% | 18% |
| **Other or unknown** | 0% | 2% |
| **Baseline characteristics (during ≤1 year index date)** | | |
| **Age at index date** | 68.5 (8.5) [61.8; 66.2; 74.7] | 70.7 (7.7) [64.8; 70.4; 76.4] |
| **Women** | 38% | 37% |
| **Charlson's Comorbidity Index** | 1.1 (1.5) [0.0; 0.0; 2.0] | 0.6 (1.2) [0.0; 0.0; 1.0] |
| **PD related diagnoses** | | |
| **Falls** | 3% | 4% |
| **Psychosis** | 1% | 1% |
| **Follow-up characteristics (during ≤2 years following index date)** | | |
| **PD Progression** | | |
| **Dementia** | 15%† (11%) | 12%† (11%) |
| **Charlson's Comorbidity Index** | 1.4 (1.9) [0.0; 1.0; 2.0] | 1.1 (1.8) [0.0; 0.0; 2.0] |

† 15% and 12% are Kaplan-Mayer estimators in MarketScan and Explorys resp., which adjust for censoring.

**Table S5. Zolpidem cohort characteristics.** Shown are the mean, standard deviation (in parentheses), and the first, second (median), and third quartile (in brackets).

| | **MarketScan** | **Explorys** |
|---|---|---|
| **Patient count** | 847 | 1,828 |
| **Patient timeline [years]** | | |
| **Total** | 4.0 (1.1) [3.0; 4.1; 5.0] | 11.4 (5.1) [7.2; 11.1; 15.2] |
| **Before index date** | 2.2 (1.0) [1.4; 2.0; 2.8] | 7.5 (4.7) [3.6; 6.7; 10.7] |
| **After index date** | 1.8 (1.1) [0.9; 1.6; 2.6] | 3.9 (2.6) [1.8; 3.4; 5.4] |
| **No. of unique prescribed drugs** | 25.1 (12.7) [16.0; 23.0; 32.0] | 40.3 (25.8) [20.0; 35.0; 55.0] |
| **Insurance** | | |
| **Medicare, Medicaid, other public** | 77% | 83% |
| **Commercial, private only** | 23% | 9% |
| **Other or unknown** | 0% | 8% |
| **Baseline characteristics (during ≤1 year before index date)** | | |
| **Age at index date** | 71.4 (10.1) [62.6; 70.0; 79.5] | 73.9 (7.8) [67.8; 74.8; 80.2] |
| **Women** | 43% | 45% |
| **Charlson's Comorbidity Index** | 2.1 (2.3) [0.0; 1.0; 3.0] | 1.7 (2.1) [0.0; 1.0; 3.0] |
| **PD related diagnoses** | | |
| **Falls** | 8% | 11% |
| **Psychosis** | 5% | 4% |
| **Follow-up characteristics (during ≤2 years following index date)** | | |
| **PD Progression** | | |
| **Dementia** | 26%† (20%) | 23%† (21%) |
| **Charlson's Comorbidity Index** | 2.8 (2.9) [0.0; 2.0; 4.0] | 3.0 (3.0) [1.0; 2.0; 5.0] |

† 26% and 23% are Kaplan-Mayer estimators in MarketScan and Explorys resp., which adjust for censoring.

**Table S6. N04 cohort characteristics.** Shown are the mean, standard deviation (in parentheses), and the first, second (median), and third quartile (in brackets).

| | **MarketScan** | **Explorys** |
|---|---|---|
| **Patient count** | 10,289 | 12,408 |
| **Patient timeline [years]** | | |
| **Total** | 4.1 (1.0) [3.1; 4.8; 5.0] | 10.9 (5.1) [6.8; 10.5; 14.6] |
| **Before index date** | 2.5 (1.1) [1.6; 2.3; 3.3] | 7.5 (4.7) [3.6; 6.5; 10.6] |
| **After index date** | 1.6 (1.0) [0.8; 1.4; 2.4] | 3.5 (2.6) [1.5; 2.8; 4.8] |
| **No. of unique prescribed drugs** | 17.4 (10.4) [10.0; 16.0; 23.0] | 18.3 (17.8) [5.0; 13.0; 25.0] |
| **Insurance** | | |
| **Medicare, Medicaid, other public** | 79% | 82% |
| **Commercial, private only** | 21% | 12% |
| **Other or unknown** | 0% | 6% |
| **Baseline characteristics (during ≤1 year before index date)** | | |
| **Age at first diagnosis** | 73.3 (9.8) [64.6; 73.6; 81.1] | 73.7 (7.9) [67.9; 74.5; 79.9] |
| **Women** | 42% | 44% |
| **Charlson's Comorbidity Index** | 1.7 (2.0) [0.0; 1.0; 3.0] | 0.8 (1.3) [0.0; 0.0; 1.0] |
| **PD related diagnoses** | | |
| **Falls** | 5% | 4% |
| **Psychosis** | 3% | 1% |
| **Follow-up characteristics (during ≤2 years following index date)** | | |
| **PD progression** | | |
| **Dementia** | 28%† (21%) | 19%† (17%) |
| **Charlson's Comorbidity Index** | 2.2 (2.5) [0.0; 1.0; 3.0] | 1.6 (2.1) [0.0; 1.0; 2.0] |

† 28% and 19% are Kaplan-Mayer estimators in MarketScan and Explorys resp., which adjust for censoring.

**Table S7. N05 cohort characteristics.** Shown are the mean, standard deviation (in parentheses), and the first, second (median), and third quartile (in brackets).

| | **MarketScan** | **Explorys** |
|---|---|---|
| **Patient count** | 3,116 | 9,067 |
| **Patient timeline [years]** | | |
| **Total** | 4.0 (1.1) [3.0; 4.2; 5.0] | 10.4 (5.0) [6.5; 9.6; 14.1] |
| **Before index date** | 2.3 (1.0) [1.5; 2.1; 3.0] | 7.1 (4.6) [3.4; 6.1; 10.1] |
| **After index date** | 1.7 (1.1) [0.8; 1.5; 2.6] | 3.3 (2.4) [1.5; 2.8; 4.5] |
| **No. of unique prescribed drugs** | 19.4 (10.0) [12.0; 18.0; 25.0] | 24.4 (17.8) [11.0; 20.0; 33.0] |
| **Insurance** | | |
| **Medicare, Medicaid, other public** | 83% | 84% |
| **Commercial, private only** | 17% | 9% |
| **Other or unknown** | 0% | 7% |
| **Baseline characteristics (during ≤1 year before index date)** | | |
| **Age at first diagnosis** | 74.0 (9.9) [65.1; 74.4; 82.1] | (7.9) [68.9; 75.4; 80.8] |
| **Women** | 48% | 45% |
| **Charlson's Comorbidity Index** | 1.8 (2.1) [0.0; 1.0; 3.0] | 1.1 (1.7) [0.0; 0.0; 2.0] |
| **PD related diagnoses** | | |
| **Falls** | 6% | 10% |
| **Psychosis** | 7% | 4% |
| **Follow-up characteristics (during ≤2 years following index date)** | | |
| **PD progression** | | |
| **Dementia** | 38%† (30%) | 29%† (27%) |
| **Charlson's Comorbidity Index** | 2.4 (2.7) [0.0; 2.0; 4.0] | 2.3 (2.6) [0.0; 2.0; 4.0] |

† 38% and 29% are Kaplan-Mayer estimators in MarketScan and Explorys resp., which adjust for censoring.

**Table S8. Estimated effects on dementia onset for emulated RCTs involving rasagiline and its encompassing ATC classes.** For composition of treatment and control cohorts, see Table S10. Beneficial effect is highlighted in green and non-beneficial effect is highlighted in red.

| | | Control cohort | | | | | | | |
|---|---|---|---|---|---|---|---|---|---|
| | | N04BD | | N04B | | N04 | | N | |
| | | Estimated effects | | | | | | | |
| | | Weight balancing | Outcome model | Weight balancing | Outcome model | Weight balancing | Outcome model | Weight balancing | Outcome model |
| | | **MarketScan** | | | | | | | |
| Treatment cohort | Rasagiline | -0.009 | -0.06 | -0.09 | -0.07 | -0.09 | -0.07 | -0.09 | -0.07 |
| Treatment cohort | N04BD | | | -0.08 | -0.06 | -0.08 | -0.06 | -0.08 | -0.06 |
| Treatment cohort | N04B | | | | | 0.13 | -0.08 | Failed to balance cohorts | |
| Treatment cohort | N04 | | | | | | | Failed to balance cohorts | |
| | | **Explorys** | | | | | | | |
| Treatment cohort | Rasagiline | -0.06 | -0.08 | -0.09 | -0.07 | -0.08 | -0.07 | -0.09 | -0.09 |
| Treatment cohort | N04BD | | | -0.11 | -0.11 | -0.11 | -0.1 | -0.08 | -0.08 |
| Treatment cohort | N04B | | | | | Failed to balance cohorts | | -0.00013 | -0.11 |
| Treatment cohort | N04 | | | | | | | -0.01 | -0.1 |

**Table S9. Estimated effects on dementia onset for emulated RCTs involving zolpidem and its encompassing ATC classes.** For composition of treatment and control cohorts, see Table S10. Beneficial effect is highlighted in green and non-beneficial effect is highlighted in red.

| | | **Control cohort** | | | | | | | |
|---|---|---|---|---|---|---|---|---|---|
| | | N05CF | | N05C | | N05 | | N | |
| | | **Estimated effects** | | | | | | | |
| | | Weight balancing | Outcome model | Weight balancing | Outcome model | Weight balancing | Outcome model | Weight balancing | Outcome model |
| | | MarketScan | | | | | | | |
| Treatment cohort | Zolpidem | -0.03 | -0.1 | -0.13 | -0.13 | -0.12 | -0.1 | -0.09 | -0.04 |
| | N05CF | | | -0.13 | -0.12 | -0.13 | -0.12 | -0.08 | -0.05 |
| | N05C | | | | | -0.06 | -0.07 | -0.05 | -0.006 |
| | N05 | | | | | | | 0.03 | 0.02 |
| | | Explorys | | | | | | | |
| Treatment cohort | Zolpidem | Control cohort too small | | 0.01 | 0.006 | -0.08 | -0.08 | -0.02 | -0.01 |
| | N05CF | | | 0.03 | 0.01 | -0.06 | -0.07 | -0.02 | -0.01 |
| | N05C | | | | | -0.12 | -0.11 | -0.06 | -0.03 |
| | N05 | | | | | | | 0.07 | 0.02 |
